# A web-based tool for real-time adequacy assessment of kidney biopsies

**DOI:** 10.1101/2024.02.01.24302147

**Authors:** Meysam Ahangaran, Emily Sun, Khang Le, Jiawei Sun, William M. Wang, Tian Herng Tan, Lyle J. Burdine, Zeljko Dvanajscak, Clarissa A. Cassol, Shree Sharma, Vijaya B. Kolachalama

**Author notes:** <u>Corresponding author</u>: Vijaya B. Kolachalama, PhD, FAHA, 72 E. Concord Street, Evans 636, Boston, MA, USA – 02118; Twitter: vkola_lab. S.S. and V.B.K. jointly supervised this work.

## Abstract

The escalating incidence of kidney biopsies providing insufficient tissue for diagnosis poses a dual challenge, straining the healthcare system and jeopardizing patients who may require re-biopsy or face the prospect of an inaccurate diagnosis due to an unsampled disease. Here, we introduce a web-based tool that can provide real-time, quantitative assessment of kidney biopsy adequacy directly from photographs taken with a smartphone camera. The software tool was developed using a deep learning-driven automated segmentation technique, trained on a dataset comprising nephropathologist-confirmed annotations of the kidney cortex on digital biopsy images. Our framework demonstrated favorable performance in segmenting the cortex via 5-fold cross-validation (Dice coefficient: 0.788±0.130) (n=100). Offering a bedside tool for kidney biopsy adequacy assessment has the potential to provide real-time guidance to the physicians performing medical kidney biopsies, reducing the necessity for re-biopsies. Our tool can be accessed through our web-based platform: http://www.biopsyadequacy.org.

## Introduction

Kidney biopsies are indispensable for diagnosing renal parenchymal diseases in both native and transplanted organs.^1^ Traditionally, pathologists have conducted real-time assessments of biopsy adequacy using microscopes,^2^ a practice vital for accurate histopathological evaluations. However, the process faces significant challenges: primarily staffing limitations in resource-constrained settings and the increasing involvement of radiologists in performing the biopsy procedures. Many healthcare institutions grapple with the inability to have a pathologist present on site, within the biopsy suite, for immediate adequacy assessment due to constraints in cost and staffing.^3^ This shortfall has led to an escalation in kidney biopsies yielding incomplete diagnostic information due to insufficient tissue samples. Consequently, this increases the healthcare burden and poses substantial risks to patients, including the possibility of undergoing repeat biopsies or receiving potentially inaccurate diagnoses based on incomplete information.

A recent study by Nissen and colleagues has shed light on this growing concern,^4^ revealing that the rate of incomplete native renal biopsies has alarmingly increased from 2% in 2005 to 14% in 2020. This increase is notably correlated with a shift in biopsy procedures from nephrologists to radiologists,^5^ particularly in the United States. In 2018, radiologists performed 95% of these biopsies, often opting for smaller-diameter needles (18g/20g), in stark contrast to only 5% in 2005. Such a transition in procedural practice has led to a decrease in critical biopsy components like the number of glomeruli per centimeter of core biopsy and the mean core width, especially when smaller needles are employed. While nephrologists are acutely aware of the necessity for an adequate amount of cortex containing glomeruli for optimal diagnostic yield, radiologists, who deal with a wide array of procedures, may have variable understanding of this crucial aspect. Therefore, they could benefit from immediate, on-site guidance regarding biopsy adequacy. These developments underscore the need for innovative, smartphone-powered technologies that can provide real-time assessment of biopsy adequacy.^6, 7^ Such technologies can substantially reduce the risk to patients and improve the accuracy of diagnoses. Here we present a web-based software developed with a deep learning framework designed for segmenting digital kidney core biopsy images and determining the cortex percentage.

## Methods

We developed a web-based application driven by deep learning for real-time assessment of kidney biopsy adequacy. This framework was trained on a dataset of digitized core biopsy images and leveraged a publicly available image segmentation architecture, the MedSAM model,^8^ as the foundation for this deep learning framework (**Figure 1**). The biopsy cores (16G) were sourced from five fresh, unutilized deceased donor kidneys and captured using an iPhone 13 Pro camera. These images were then processed through formalin fixation, paraffin embedding, and sectioning to produce PAS-stained slides, establishing a histological correlation.

**Figure 1.**
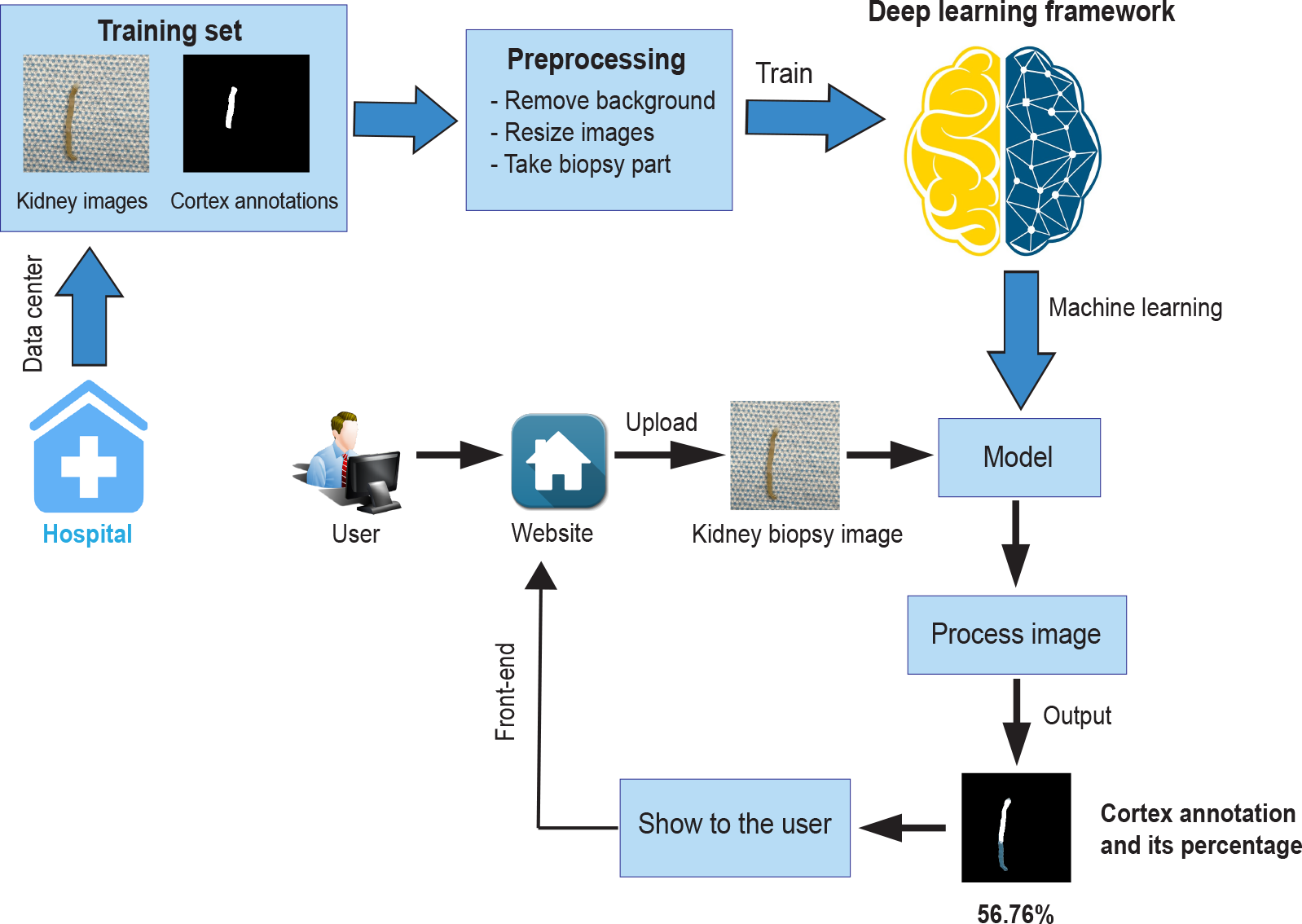
Web-based tool for kidney biopsy adequacy assessment. Our web-based tool employs a machine learning model that has been trained on a dataset of kidney cortex annotations provided by nephropathologists. It comprises both a front-end and a back-end component. First, the users are given the option to upload a kidney biopsy image via the website. After the user uploads and submits the image for analysis, the back-end server processes it. The tool generates an image with the kidney cortex highlighted and calculates the percentage of cortex. The result is presented to the user, including the model-processed image showcasing the cortex area and the associated percentage value.

Our team of expert nephropathologists, boasting over 21 years of combined experience (Mean±std: 7±3.27 years), collaboratively annotated the images. We applied the MedSAM model to 100 resized biopsy images for segmentation analysis, employing a 5-fold cross-validation method for internal validation. This involved dividing the data into training (80%) and test (20%) sets, ensuring a rigorous evaluation of the model’s performance and generalizability with Intersection over Union (IoU) and Dice coefficient metrics.

We engineered a user-centric web application that allows for the uploading, cropping, and analyzing of biopsy images. This platform developed using JavaScript for the front-end and Django for the back end, is supported by a robust computing infrastructure. More details on the methodology are presented in the **Supplement**.

## Results

We evaluated our model’s performance using 5-fold cross-validation and applied two key metrics - Intersection over Union (IoU) and Dice coefficient - which are standard in medical image segmentation evaluation. The model demonstrated impressive performance across a set of 100 kidney biopsy images, achieving an IoU score of 0.665 (±0.144) and a Dice coefficient of 0.788 (±0.130). These metrics gauge the similarity between the expert-annotated and model-predicted cortex segments of kidney images, with their values ranging from 0 (indicating no overlap) to 1 (indicating perfect annotation).

In **Figure 2**, we illustrated the model’s performance across five representative kidney biopsy images. This includes both the actual cortex regions, identified by medical experts, and the model’s predictions, with white pixels indicating cortex areas. We also provided the actual and predicted cortex percentages for each image, representing the proportion of cortex pixels relative to the total pixels in the kidney biopsy image, on a scale from 0 to 1. Our results demonstrate that the model effectively handles kidney biopsy images with varying levels of cortex presence. Also, the model demonstrated strong reliability in accurately identifying cortex regions in kidney biopsy images, as evidenced by a high correlation between predicted and actual cortex percentages. This finding is supported statistically with a Spearman correlation coefficient of 0.724 and a highly significant p-value of 1.632e-17.

**Figure 2.**
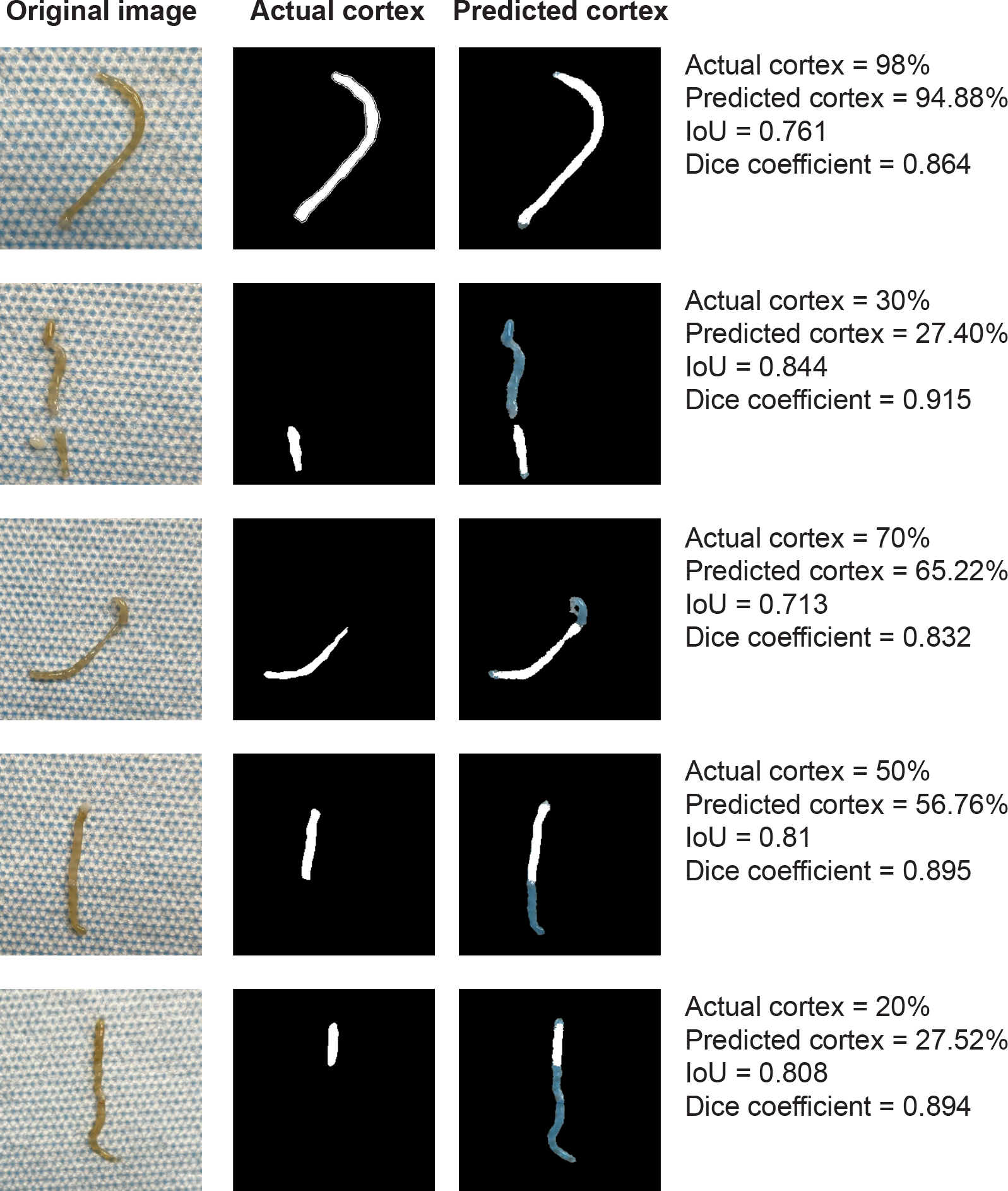
Sample kidney biopsy images along with their annotations and model predictions. The figure includes five representative cases, each illustrating the actual cortex regions, as determined by nephropathologists, and the corresponding cortex regions as predicted by our model. For each case, two critical pieces of information are provided: the actual cortex percentage and the predicted cortex percentage. These percentages indicate the proportion of cortex pixels relative to the total pixel count in each kidney biopsy image and are scaled on a range from 0 to 1. To quantitatively assess the model’s accuracy in cortex annotation for each kidney biopsy image, we employed two essential segmentation metrics: the Intersection over Union (IoU) and the Dice coefficient. These metrics measure the similarity between the expert-annotated and model-predicted cortex regions, with a value range from 0 (indicating no similarity) to 1 (indicating perfect match in annotation).

## Discussion

Our study aimed to develop a tool that supported practitioners conducting biopsies for renal disease investigations by providing real-time estimations of cortex presence in tissue cores, thereby enhancing biopsy adequacy. Utilizing a deep learning framework augmented by expert annotations on digitized biopsy images, we established a proof-of-principle that demonstrated the feasibility of our approach. The resultant web-based tool represents a practical application of this technology, offering immediate assistance to practitioners by accurately assessing biopsy tissue adequacy in real time.

Our tool offers a user-friendly interface that is readily accessible through any modern web browser such as Chrome or Firefox. To ensure security and manageability, the tool requires user registration. New users are guided to create an account with verifiable credentials, while returning users can simply log in with their existing credentials. Once registered and logged in, researchers have the capability to upload digitized images of biopsy cores in common formats such as JPG, PNG, and TIFF. After uploading an image, users are provided with a feature to delineate the region of interest, specifically the area of the biopsy within the image. This functionality enables users to crop the biopsy area, excluding extraneous background elements, thereby focusing the analysis on the biopsy itself. This not only improves the quality of the output but also enhances user engagement. Upon confirming and submitting the uploaded image for analysis, the platform generates two outputs: (1) An annotated image showing the cortex area within the core biopsy. (2) A calculated percentage representing the cortex area in relation to the entire core biopsy image. Our software is adept at handling various scenarios, including digitized biopsy images that may entirely lack cortex or contain the kidney cortex in full. In instances where the estimated cortex percentage is either 0 or 100, the software’s response varies accordingly. If the predicted percentage is 100%, the software will display the cortex percentage, and the cortex visualization will encompass the entire biopsy image in the output. Conversely, if the estimation is 0%, then no cortex will be highlighted on the biopsy image. In this scenario, the displayed output will resemble the original image uploaded by the user, with no discernible cortex area. After seeing the displayed percentage of the cortex, the adequacy can be estimated by the user depending on their needs. For optimal diagnostic accuracy, we recommend that a biopsy is considered adequate when the model’s estimation of the cortex area reaches or exceeds 70%.

For capturing optimal biopsy images, we recommend users to utilize the camera of a modern smartphone. Modern devices capable of capturing high-resolution photographs are crucial for precise analysis. For optimal results, images should be taken against a light, uniform background, ensuring effective processing and reliable analytical outcomes. Users are encouraged to upload multiple images of the same biopsy sample for a more comprehensive adequacy estimate. While our tool currently supports only single-image uploads, manually averaging the values from multiple uploads can offer a more accurate and reliable assessment, despite being more time-consuming.

Our study has a few limitations. We developed our model on a set of consensus annotations performed by a team of nephropathologists practicing in the United States. For the tool to be generalizable across practices worldwide, it would be important to build the model on a more diverse group of experts. We plan to refine our model, initially developed with images from non-perfused kidneys where distinguishing cortex and medulla is challenging, by using images from perfused kidneys to enhance its accuracy for real-world applications. Our digital image capture protocol utilized a single smartphone, a clean background, and a set distance between the biopsy and camera, with plans to test the tool’s effectiveness on varying backgrounds, distances, and imaging technologies in future studies.

In conclusion, our web-based tool offers an effective means for real-time, quantitative evaluations of kidney biopsy adequacy, proving especially beneficial in settings with limited resources and no immediate access to pathologists. By utilizing a simple photograph and internet access, clinicians can quickly ascertain the adequacy of a biopsy, facilitating prompt decisions on whether further samples are needed. While additional research is necessary to gauge its full effects on patient care and healthcare systems, this innovation represents a significant step forward in incorporating AI technology into kidney disease management.

## Data Availability

All data produced in the present study are available upon reasonable request to the authors.

## Data sharing statement

Biopsy image data will be made available on request. Computer scripts and manuals are made available on GitHub: https://github.com/vkola-lab/ki2024.

## Acknowledgments

This work was supported by the NIH SBIR phase 1 grant (R43-DK134273). V.B.K. also acknowledges support from the following sources: the Karen Toffler Charitable Trust, the American Heart Association (20SFRN35460031), the National Institutes of Health (RF1-AG062109, R01-HL159620, and R21-CA253498), and the National Institute on Aging’s Artificial Intelligence and Technology Collaboratories (AITC) for Aging Research program.

## Methods

### Study population

We acquired 5 fresh, unutilized deceased donor kidneys from the University of Arkansas for Medical Sciences (UAMS) in Little Rock. From these kidneys, we performed a total of 747 core biopsies using a 16G needle. For the purposes of our study, we selected the first 500 images. Each biopsy core was photographed using a handheld iPhone 13 Pro camera. To ensure a clear focus on the kidney core, the camera was not mounted on a tripod but held approximately 2 inches from the tissue by hand. The biopsy cores were free of blood, and the background for each image was the standard absorbing paper typically used in pathology laboratories. Following the initial photography, each biopsy core underwent formalin fixation, paraffin embedding, and sectioning. From this process, we produced a single Periodic Acid–Schiff (PAS) stained slide for each specimen. These slides were then scanned at 20x magnification using our Olympus VS200 whole slide imaging system. The primary purpose of these high-resolution images was to establish a correlation between the histology of the PAS-stained slides and the gross morphology observed in the core biopsies. This comprehensive approach was designed to enhance our understanding of the relationship between the microscopic structure and the gross anatomical features of the kidney tissues.

### Expert-driven annotations

The initial phase of our study involved uploading core biopsy images to PixelView (deepPath, Inc., Boston, MA), a web-based tool for digital pathology analysis. Three nephropathologists collaboratively annotated the first 20 images, establishing a consensus on the annotations. Based on this consensus, one nephropathologist proceeded to annotate the subsequent 80 images. The first set of 20 images underwent detailed annotation, focusing on distinct anatomical areas: the cortex, medulla, and fibrofatty tissue. Histology slides served as essential references during this process. However, the annotation task presented several challenges. A primary issue was the difficulty in distinguishing between the cortex and medulla in the gross images due to subtle color gradations. This problem was more noticeable in biopsies from deceased donors, where the kidney cores often had a shiny appearance, making the distinction less clear compared to those from fresh biopsies. Additionally, the transparency of some tissues allowed the background to be visible, complicating the distinction between cortex and medulla areas. A similar challenge was faced in differentiating the capsule from adjacent fibroconnective tissue. Considering these challenges and our goal of determining the biopsy’s adequacy with absolute specificity, we decided to focus solely on annotating the cortex in the remaining images. This decision was informed by the clinical importance of the cortex in biopsy evaluations, as it is the primary area of interest during a biopsy. Therefore, it was concluded that providing percentages for all biopsy components was not essential for our analysis. A single nephropathologist, guided by the histology images, subsequently annotated only the cortex in the remaining 80 gross biopsy core images. In total, the annotations of all 100 images, including those from the initial collaborative effort and the subsequent individual annotations, were utilized for the development of our AI model. This approach in the annotation process ensured consistency, laying a foundation for the AI model’s reliability in analyzing and interpreting kidney biopsy images.

### Machine learning framework

We developed a deep learning framework for segmenting medical images using 100 gross biopsy images and their corresponding cortex annotations. This framework is based on the MedSAM model,^1^ an advanced version of the Segment Anything Model (SAM) tailored for medical imagery.^2^ MedSAM has been trained on a comprehensive dataset of 1,570,263 image-mask pairs across 10 imaging modalities, demonstrating superior precision in medical image segmentation tasks. Our process began by resizing each gross biopsy image and its annotation to a uniform size of 256x256 pixels. These resized image-annotation pairs were then fed into the MedSAM framework for segmentation analysis. For internal validation, we employed a 5-fold cross-validation method. This involved dividing the data into training (80%) and test (20%) sets, training the model on the training set, and then evaluating it with the test set. This cycle was repeated five times to ensure an unbiased evaluation of the model’s performance and generalizability. Using the pre-trained MedSAM model as a foundation, we could exploit its extensive learning capabilities, resulting in more precise and efficient segmentation that meets the specific requirements of biopsy image analysis.

### Performance metrics

We assessed the performance of the model by employing metrics such as Intersection over Union (IoU) and Dice coefficient to measure the congruence between the kidney cortex predictions generated by the model and expert annotations. These two metrics are used to evaluate the performance of object detection models by calculating the similarity between predicted segmented object and the ground truth object. Biopsy adequacy, defined as a percentage representing the cortex area relative to the overall biopsy area, was used for evaluation.

### Software development

We engineered an advanced web-based application utilizing the robust capabilities of JavaScript and the Django framework,^3^ which are integral to its front-end and back-end architecture, respectively. On the front-end, the website offers a user-centric interface where users can upload high-resolution images of kidney biopsy samples. The tool features an advanced cropping function that enables precise isolation of the main biopsy area, enhancing the accuracy and efficiency of the analysis by focusing on the most relevant section of the image. The front-end was developed using JavaScript, which is a versatile and widely used programming language, essential for adding interactive elements to web pages, and is supported by all modern web browsers, making it a foundational tool for web development. Once the user completes the cropping, the image is seamlessly transferred to the back-end server, built on the Django framework. Django, a high-level Python web framework, is renowned for its rapid development capabilities and strong emphasis on security, offering a built-in admin panel, excellent documentation, and a scalable architecture that makes it suitable for a wide range of web applications.

The server employs the machine learning framework (described above) to meticulously analyze the kidney biopsy image. This analysis involves identifying and highlighting the cortex parts of the kidney, which are crucial for assessing various renal conditions. The back-end server’s algorithm is designed to accurately calculate the proportion of the cortex in the biopsy image. Following the analysis, the server generates a detailed output image that visually highlights the cortex areas for easy interpretation. Alongside the visual output, the tool provides a quantitative assessment, displaying a numerical value that indicates the percentage of cortex regions relative to the entire area of the biopsy image. This quantitative analysis is essential for medical professionals in evaluating the extent of certain kidney conditions, such as glomerular diseases, which may affect the cortex region.

### Computing infrastructure

We used PyTorch (v1.9.0) and a NVIDIA 2080Ti graphics card with 11 GB memory on a GPU workstation to implement the model. The training speed was about 0.05 iterations/s, and it took 3 hours to reach convergence. The inference time for a standard biopsy image of size 1 Mb is within a minute.

